# *De novo* identification and visualization of important cell populations for classic Hodgkin lymphoma using flow cytometry and machine learning

**DOI:** 10.1101/2020.12.18.20248526

**Authors:** Paul D. Simonson, Yue Wu, David Wu, Jonathan R. Fromm, Aaron Y. Lee

## Abstract

**Objectives:** Automated classification of flow cytometry data has the potential to reduce errors and accelerate flow cytometry interpretation. We desired a machine learning approach that is accurate, intuitively easy to understand, and highlights the cells that are most important in the algorithm’s prediction for a given case.

**Methods:** We developed an ensemble of convolutional neural networks (CNNs) for classification and visualization of impactful cell populations in detecting classic Hodgkin lymphoma, using two-dimensional (2D) histograms. Data from 977 and 245 clinical flow cytometry cases were used for training and testing, respectively. 78 non-gated 2D histograms were created per flow cytometry file. SHAP values were calculated to determine the most impactful 2D histograms and regions within the histograms. The SHAP values from all 78 histograms were then projected back to the original cells data for gating and visualization using standard flow cytometry software.

**Results:** The algorithm achieved 67.7% recall (sensitivity), 82.4 % precision, and 0.92 AUROC. Visualization of the important cell populations in making individual predictions demonstrated correlations with known biology.

**Conclusions:** The method presented enables model explainability while highlighting important cell populations in individual flow cytometry specimens, with potential applications in both diagnosis and discovery of previously overlooked key cell populations.

## Introduction

Classic Hodgkin lymphoma (cHL) is a B cell lymphoma composed of neoplastic Hodgkin cells and multinucleated Reed-Sternberg cells in a background of non-neoplastic, reactive immune cells. The disease often affects younger individuals and commonly demonstrates mediastinal involvement^1,2^. Given the sensitive location of involvement and high frequency of non-neoplastic causes of lymphadenopathy, minimally invasive, small needle core biopsies and fine needle aspirations are often the first biopsies performed. The small biopsy sizes and relatively sparse distribution of neoplastic cells reduce the sensitivity for detection of classic Hodgkin lymphoma by morphology^3,4^. Increased sensitivity can be achieved by adding flow cytometry to the armamentarium, and flow cytometry methods for the diagnosis of cHL have been demonstrated in the clinical laboratory environment^5,6,7^.

Despite this, flow cytometry for classic Hodgkin lymphoma is not widely available, possibly due to the perceived high-level of expertise required for interpretation^8^. The relative infrequency of positive samples and lack of interpretation experience makes validation of the assay challenging for many labs. Application of machine learning to flow cytometry data has the potential to reduce data interpretation subjectivity and increase accuracy in the interpretation of classic Hodgkin lymphoma flow cytometry data^9,10^. In a prior report^8^, random forest and support vector machine classifiers were used to classify classic Hodgkin lymphoma flow cytometry data. Various approaches to explain the inner workings of the models were attempted, including random forest tree analysis, principal component analysis, and accuracy comparisons when data for various antibodies were excluded. These attempts at model explanation suggested that cell populations other than Hodgkin cells (including CD5-positive cells), were important in making the classification prediction. However, details regarding predictive cell populations were lacking. Furthermore, the approaches were incapable of determining what cell populations were most impactful for making predictions for *individual* flow cytometry cases. Visualization of impactful cell populations for individual cases was therefore also impossible.

Recent work in machine learning explainability has resulted in new tools for understanding the general features that are important for a model, as well as tools for identifying the impact of individual data elements on individual predictions^10,11,12,13,14^. One such tool, the calculation of SHAP (SHapley Additive exPlanations) values, allows the identification of impactful data features on predictions, both generally and for individual predictions, by estimating the additive impact of each input data point in arriving at a classification prediction^11,13^.

Building on prior work, we sought to leverage the spatial information in histograms and build a machine learning framework with the explicit goal of interpretability and identification of important classes of cells from flow cytometry data.

## Materials and Methods

### Patient Samples

In total, 1222 samples were analyzed using a nine-color classic Hodgkin lymphoma-specific flow cytometry panel and used in accordance with University’s institutional review board (IRB) approval. Given the retrospective use of the data is for clinical laboratory test quality and operations improvement and there is minimal risk for patient harm, patient written consent was deemed unnecessary and was therefore waived by the IRB of the University. All methods were carried out in accordance with relevant guidelines, regulations, and approval by the IRB of the University. Cases from 2010 to 2019 were used. Interpretations of data in clinical reports were reviewed in order to create annotations to train a binary classifier. Cases that were reported as “consistent with”, “highly suggestive of”, or other similar language, for classic Hodgkin lymphoma were annotated as positive (N = 149). Cases that were interpreted as “suspicious for” classic Hodgkin lymphoma or similar language were also annotated as positive (N = 175), though a further review of the medical record was performed for these cases in order to determine the predictive accuracy of the interpretation. 70 cases were thus reviewed using concurrent/follow-up histology-based diagnostic results. In 3 cases, the result was negative for cHL, and those three cases were re-annotated as negative for training purposes (leaving N = 172). A small number of cases were reported as consistent with the Hodgkin component of grey zone lymphoma; these were also annotated as positive for the purposes of our classifier. Total positive cases were then 321, and total negative cases were 921. 241 cases were interpreted as consistent with or suspicious for a neoplasm other than cHL. 123 cases (not including those consistent with or suspicious for neoplasm) had some language included that suggested there was something suboptimal regarding the submitted sample (few cells, degradation, etc.); these cases were *not* excluded from the training or test data sets. Total training cases were 977 and testing cases were 245. Training cases were further subdivided into training set #1 (654) and training set #2 (323).

### Flow Cytometry and Manual Classification

Using a method previously described by Fromm et al.^5,6^, cases were evaluated on a modified four-laser, 10-color Becton Dickinson (Franklin Lakes, NJ) LSRII flow cytometer using the following laser-fluorochrome combinations: (1) a 405 nm violet laser (one color) exciting Pacific blue (PB); (2) a 488 nm blue laser exciting fluorescein isothiocyanate (FITC), phycoerythrin (PE), PE-Texas red (ECD/PE-TR), PE-Cy5.5, and PE-Cy7; and (3) a 633 nm red laser (three colors) exciting allophycocyanin (APC), APC-Alexa Fluor 700 (APC-A700), and APC-Cy7. The specific fluorescently labeled antibodies consisted of CD95-PB, CD64-FITC, CD30-PE, CD5-ECD, CD40-PECy5.5, CD20-PECy7, CD15-APC, CD71-APC-A700, and CD45-APC-Cy7, which, in addition to four light scatter properties (forward and side scatter, area and height), generated a total of thirteen dimensions of data. Typically, 0.5 to 1 million events were collected per case though cases with as few as 20,000 cells were also seen, and sequential gating was used to select for discrete populations of Hodgkin/Reed Sternberg (HRS) cells as previously described ^5,6^.

### Construction of 2D Histograms

Software was written using Python 3.7^15^ along with external python modules that included fcsparser, sklearn^16^, tensorflow (version 2.0, with included keras framework), matplotlib ^17^, seaborn, numpy ^18^, pandas, and shap^12,13^. Flow cytometry FCS files were imported via fcsparser and compensated for fluorescence channel cross talk. All values, except for forward scatter values, that were < 1.0 were then converted to 1.0, and the logarithm base 10 was applied. These values were then divided by 12.5, which achieved approximate normalization. Forward scatter values were rescaled on a linear scale with modified offset and approximate normalization to make better use of the range [0,1]. Two-dimensional (2D) histograms were then calculated for each possible pairwise combination of parameters (9 fluorescence parameters and 4 scattering parameters, making 13 total parameters, with 78 possible pairwise combinations). The 2D histograms were modified by adding 1.0 to each value, then applying a logarithm (base 10) function. Each histogram was then scaled by its maximum bin value in order to normalize each histogram individually. The histogram binning resolution was 50 × 50 on the interval [0, 1]. The data were divided into training and test data sets (80:20 ratio).

### EnsembleCNN Classifier

A CNN classifier was created to correspond to each previously constructed 2D histogram (see above); 78 were created in total. Each CNN classifier was constructed as sequential layer models using the keras framework in tensorflow 2.0 (Fig. 1). The prediction probabilities of cHL positivity for each CNN were passed as an ensemble of parameters to a random forest classifier, which gave the final prediction for each flow cytometry case (Fig. 1). Training set #1 (see Patient Samples above) was used to train the CNNs, and training set #2 was used for training the ensemble random forest classifier. In order to achieve approximately class-balanced data, the positive cases in training set #1 were duplicated twice. The CNNs were trained by using batch sizes of 200 from training set #1 for 200 epochs, with L2 regularization and parameter dropout layers to avoid overfitting. Evaluation for overfitting by CNNs was performed by using training set #2 as a validation set, and plots of training and validation data loss and accuracy versus epoch were inspected for overfitting.

**Figure 1.**
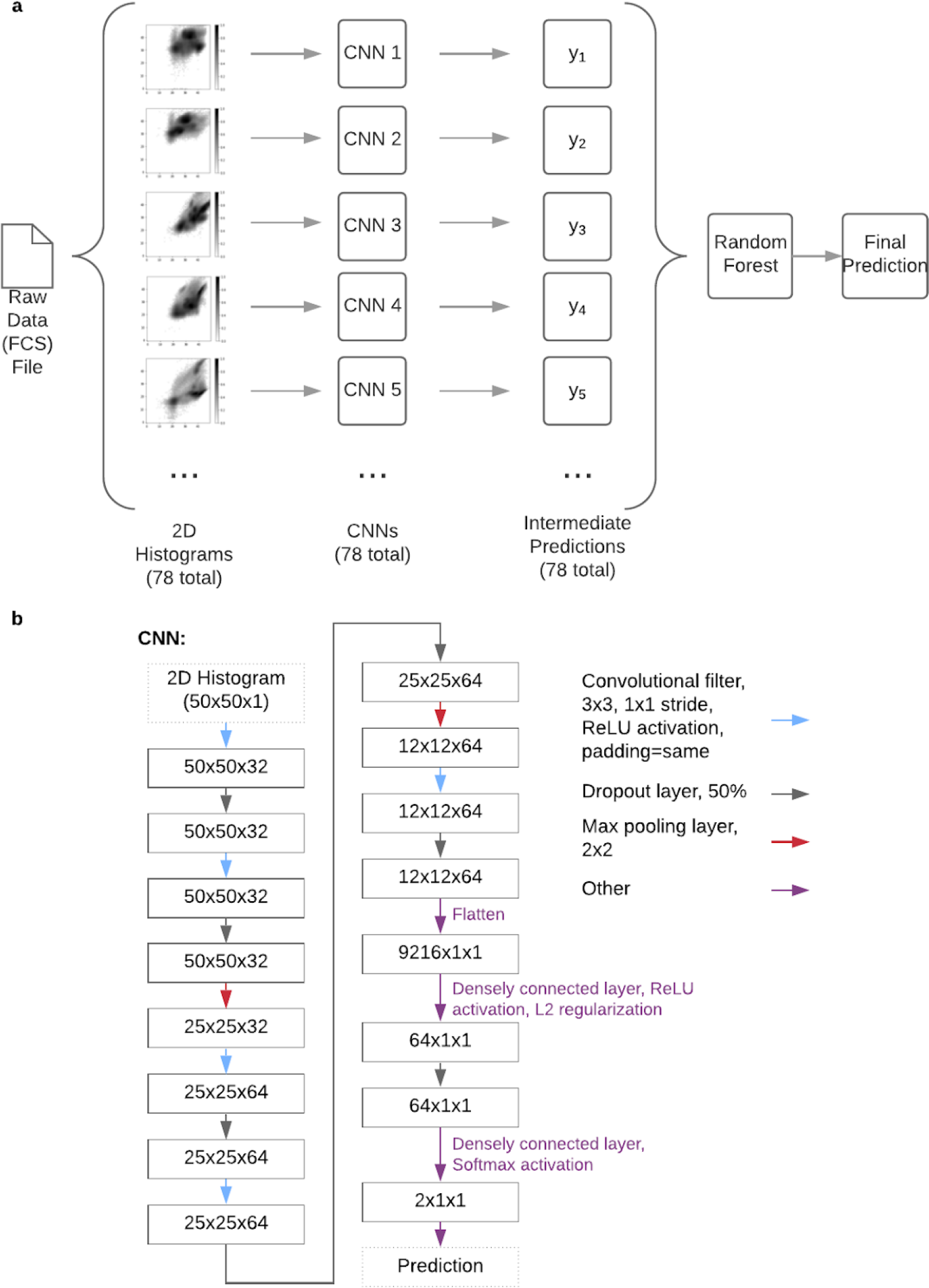
Classification model architecture. **a**. Overview of the CNN ensemble classifier. A flow cytometry data file is used to construct 2D histograms, which are then passed as inputs to an ensemble of corresponding CNNs. The outputs are passed to an integrating random forest classifier, which produces a final prediction score. **b**. Architecture used for individual CNNs. Colored arrows indicate operations performed in the various layers.

The trained CNNs were applied to training set #2 in order to produce the inputs for training the random forest classifier. The ensemble random forest classifier was implemented using sklearn’s RandomForestClassifier class with n_estimators=1000, max_leaf_nodes=32, and balanced class weighting.

### Evaluation Metrics, Including SHAP Values

A confusion matrix was generated for the test predictions and predicted classes using sklearn. R, RStudio, and the precrec library^19^ were used to plot the ROC curve and precision-recall curve seen in Fig. 3. Confidence intervals were generated by pooling the training and test sets data, then performing 5-fold crossvalidation, with the resulting predictions used to estimate the 95% confidence intervals using the precrec library.

SHAP values were calculated using the shap python module^13^ in order to estimate the impact of CNN outputs on the final prediction produced by the random forest classifier. SHAP values were also calculated for individual bins within the 2D histograms. In addition, the SHAP values from the 2D histograms were projected back to the individual cells in the original FCS files by summing the SHAP values from every bin within which a particular cell of the given FCS file was found. Three tallies for each cell were calculated: total SHAP value, positive SHAP values (only add the bin’s SHAP value if it is positive), and negative SHAP values (only add the value if it is negative). A new FCS file was then created by adding the tallies as new parameters for each cell. The new FCS file was then used for gating and visualization purposes in clinical flow cytometry analysis software (WoodList 3.1.3). See Fig. 7 for example output.

## Results

Our machine learning framework is composed of an ensemble of CNNs that each make independent predictions based on two-dimensional (2D) histograms as inputs. The predictions are passed to a random forest classifier for final prediction. The ensemble of CNN classifiers (EnsembleCNN) approach is analogous to manual diagnostic approaches already used by hematopathologists and others, and is therefore intuitive to understand. We use SHAP values to highlight the impactful 2D histograms and regions within 2D histograms.

### EnsembleCNN Classifier Results

Training of the CNNs (Figs. 1 and S1) demonstrated that the majority achieved >70% validation accuracy. Individual CNN predictions also generally correlated among themselves for a given flow cytometry specimen, as is well demonstrated in the hierarchical clustering dendrogram plot shown in Fig. 2. These findings suggest that detectable, predictive information is in fact evident to the CNNs in most of the non-gated 2D histograms (see below). The EnsembleCNN classifier resulted in a classification accuracy of 88.2%, a precision of 82.4%, a recall (sensitivity) of 67.7%, and an F1 score of 74.3%. (The full confusion matrix is available in Supplemental Table ST1.) The area under the receiver operating characteristic (AUROC) curve was 0.92 (Fig. 3). Subsequent 5-fold cross validation demonstrated similar results, with resulting generation of 95% confidence intervals in the ROC and precision-recall curves plotted in Fig. 3.

**Figure 2.**
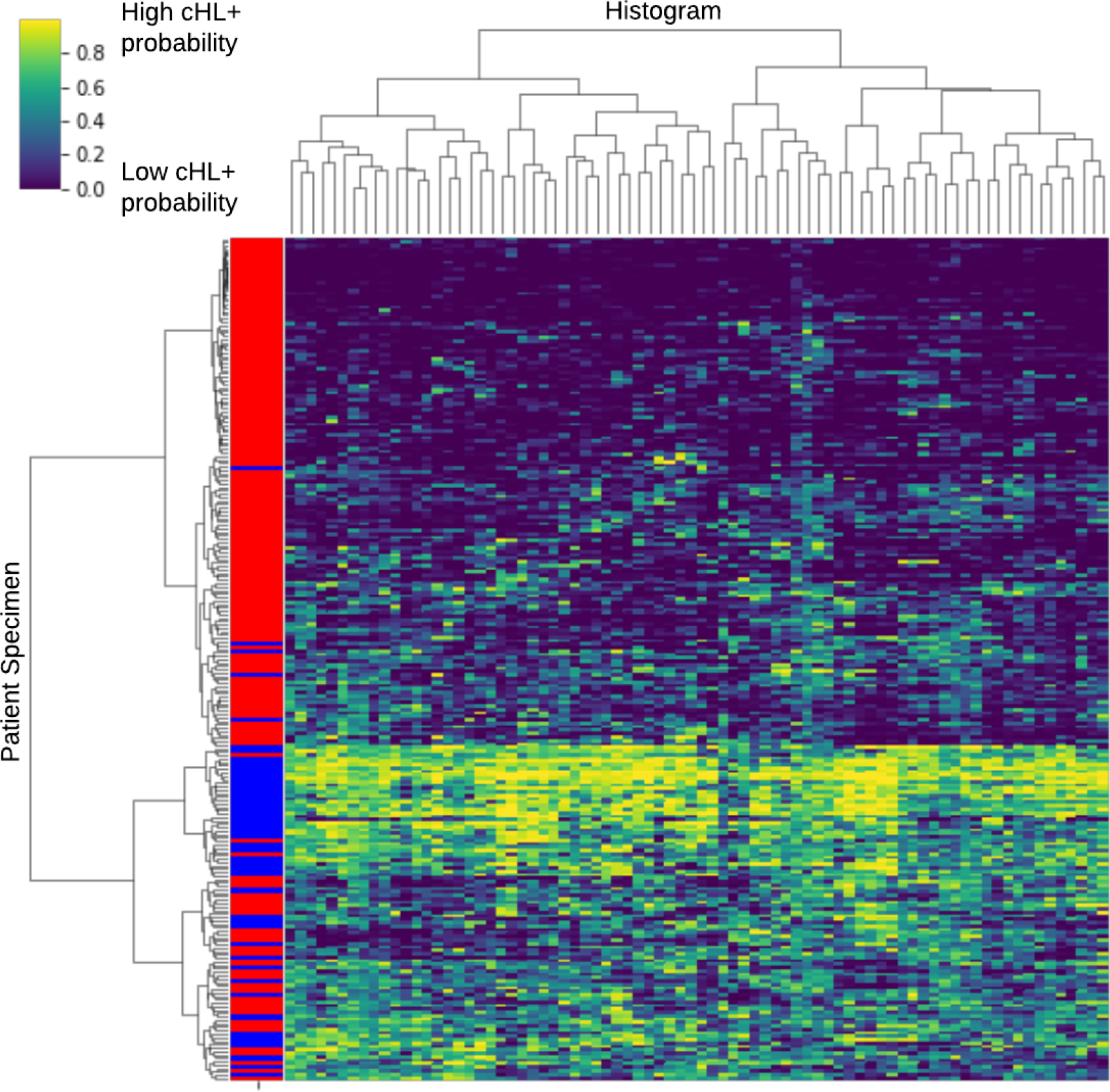
Hierarchical dendrogram of class predictions produced by CNNs applied to the test data set. Rows correspond to individual flow cytometry cases, and each has 78 class predictions, one for each of the CNNs. Row labels (left side of plot) correspond to data labels (blue = positive for cHL, red = negative). The plot demonstrates that for a single row, the predictions of the various CNNs tend to correlate amongst themselves and also with the annotated data labels.

**Figure 3.**
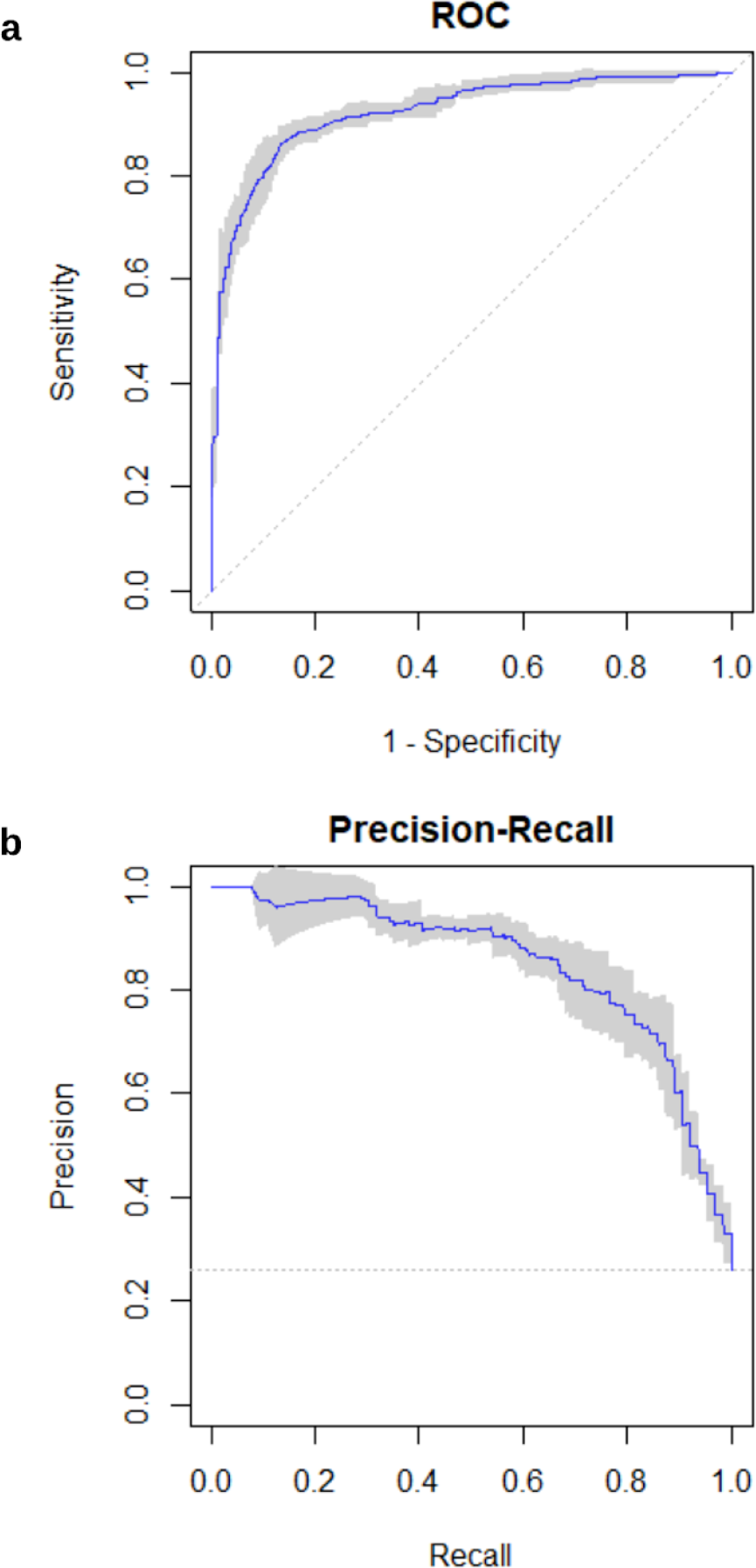
ROC curves and precision-recall curves for the CNN ensemble classifier. **a**. ROC curve, constructed using class predictions for reserved test data. Area under the curve = 0.92. **b**. Precision-recall curve. Shaded areas represent estimated 95% confidence intervals.

### SHAP values identify the most impactful 2D histograms

Given the modular framework of our EnsembleCNN classifier, we can calculate SHAP values for the various components to understand the contributions/impact of each part. Given that predictions are calculated for each 2D histogram and passed to an integrating random forest classifier (Fig. 1), we calculated SHAP values for each of the predictions passed to the random forest classifier, with the purpose of identifying the most impactful 2D histograms. The following five were found to be the most impactful: CD40/CD15, CD20/CD71, SSC-H/CD20, CD5/CD15, and SSC-A/CD20. The SHAP values for the CNN outputs for the top 20 histograms are shown in Fig. 4. The SHAP values not only allow the estimation of the overall importance of the different 2D histograms, but also yield additional insights into the behavior of the classifier. For example, the FSC-A (forward scatter) vs. CD30 histograms demonstrated little impact on the random forest prediction scores for low CNN prediction values. However, when the CNN predictions were high, there was a much larger impact on the overall prediction score. Two-dimensional dendrogram plots (Figs. 2, S2, and S3) also highlight patterns of correspondence between data classification, individual histogram prediction scores, and SHAP values.

**Figure 4.**
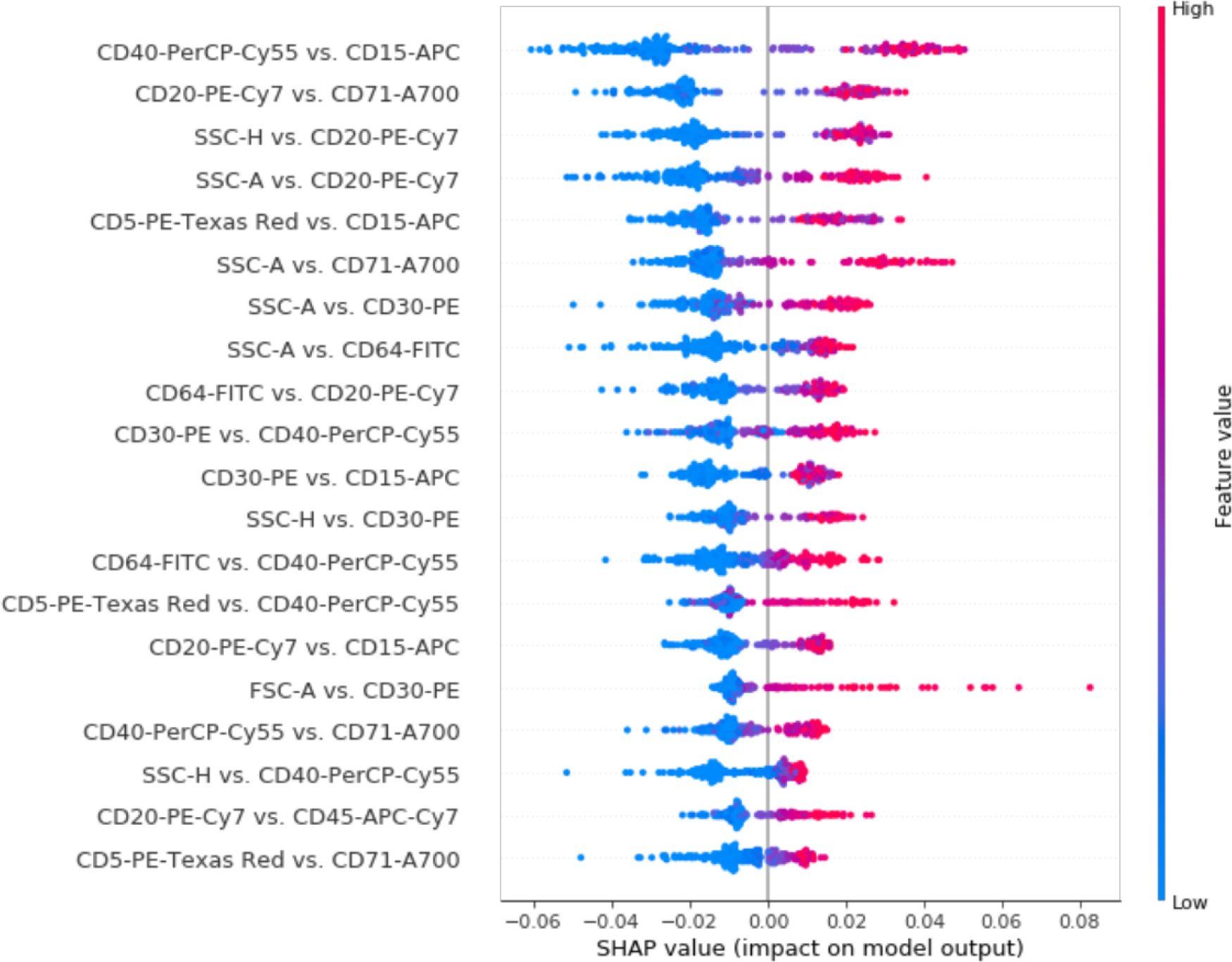
SHAP values for top 20 most impactful features for the integrating random forest classifier. Each feature is a prediction probability score (range = [0, 1]), produced by the CNNs for the corresponding 2D histograms using test set data. Dots correspond to individual flow cytometry cases in the test data set. Of interest is the strong positive impact on final prediction that the FSC-A vs. CD30-PE prediction score has when it has a high value, but little negative impact when it has a low value, for the given cases. See also Fig. S2, which presents the SHAP values for the various 2D histograms as a 2D hierarchical dendrogram.

### SHAP values highlight the most impactful bins within individual histograms

SHAP values were also calculated for individual 2D histogram bins in order to identify the regions of most impact within the 2D histograms. Example SHAP value plots for 2D histograms are shown in Figs. 5 and 6. In Fig. 5, the SHAP values are averaged over many predictions to highlight the recurrently most important parts of the 2D histograms. In Fig. 6, we plot the SHAP values specific to individual flow cytometry cases, which can be used to explain the models’ predictions on a case-by-case basis. The plotted SHAP values can be used to correlate directly with 2D histograms that pathologists and researchers are already familiar with, making for better model explainability. Most of the inspected 2D SHAP plots highlight multiple impactful regions, suggesting that in fact the presence, absence, or altered ratios of multiple cell populations play a role in the classification of classic Hodgkin lymphoma for these CNNs.

**Figure 5.**
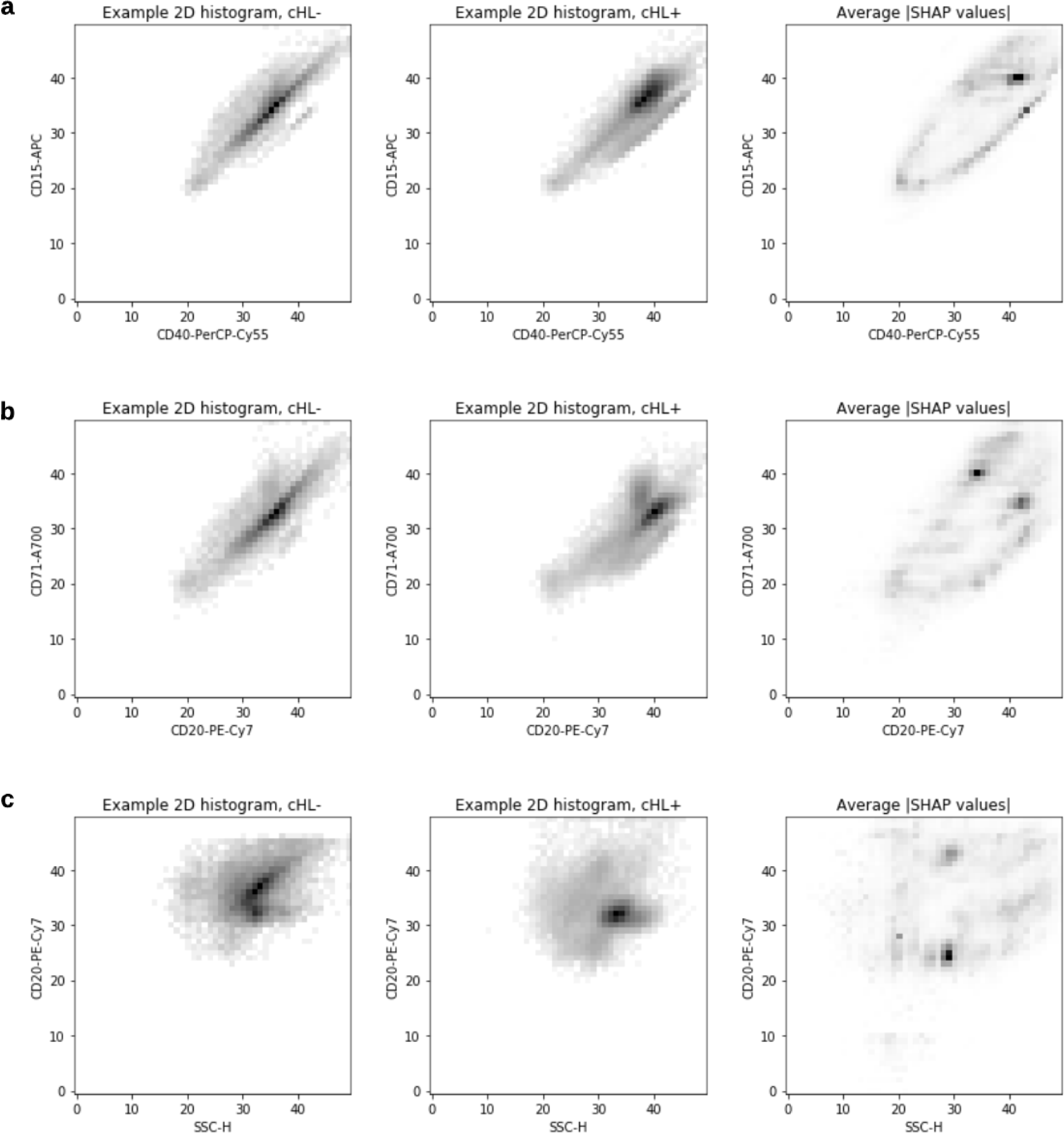
Examples of averaged (N=100) absolute values of SHAP values for 2D histograms. Axes are numbered in terms of 2D histogram bin numbers. **a**. CD40 vs. CD15. **b**. CD20 vs. CD71. **c**. SSC-H (side scatter) vs. CD20.

**Figure 6.**
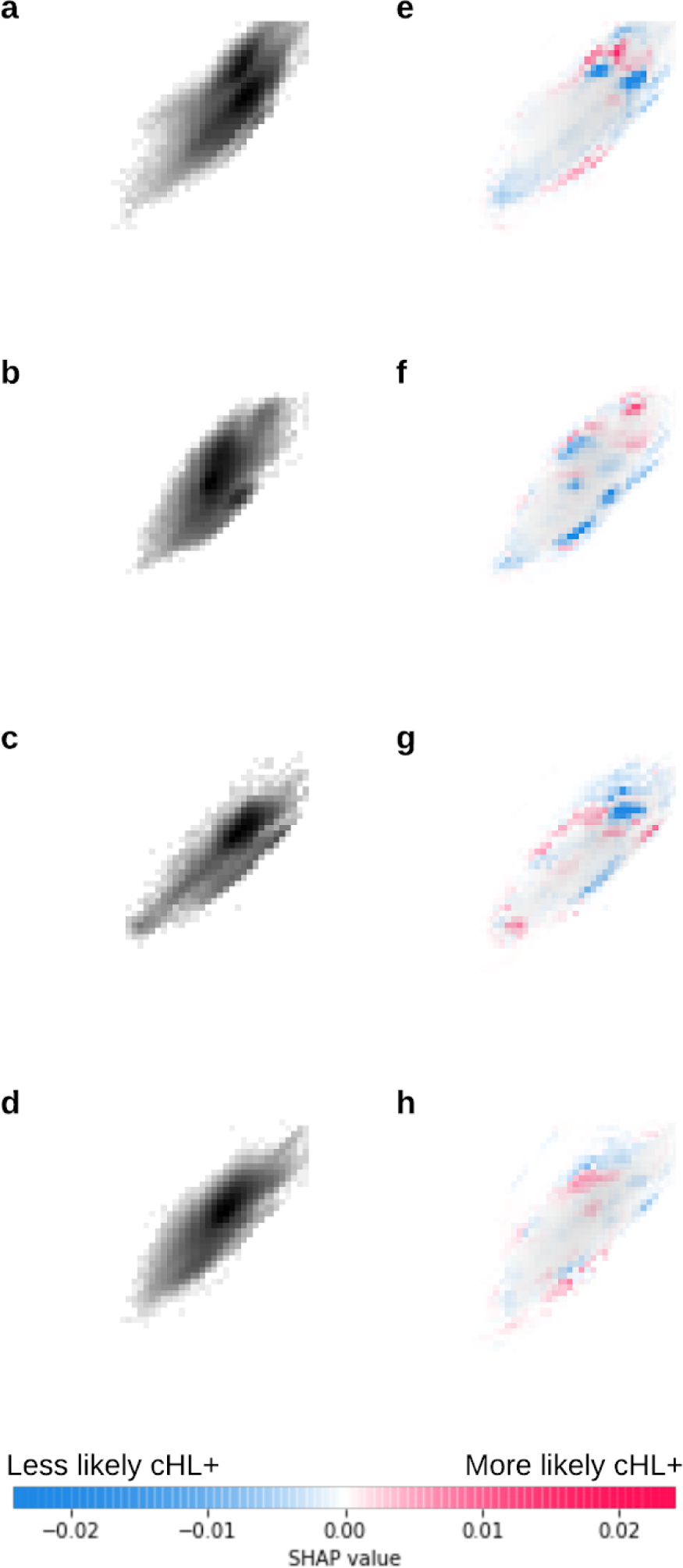
Example SHAP values plots for individual flow cytometry cases. Images **a-d** on the left are the 2D histograms for CD40 (x axes) vs. CD15 (y axes). Images **e-h** on the right show the corresponding SHAP values in red and blue, with red more predictive of Hodgkin lymphoma. A light-gray background that depicts the original histogram is also included in **e-h** for context in the images on the right. Examples were chosen from cases with predicted cHL+ probability > 0.95.

**Figure 7.**
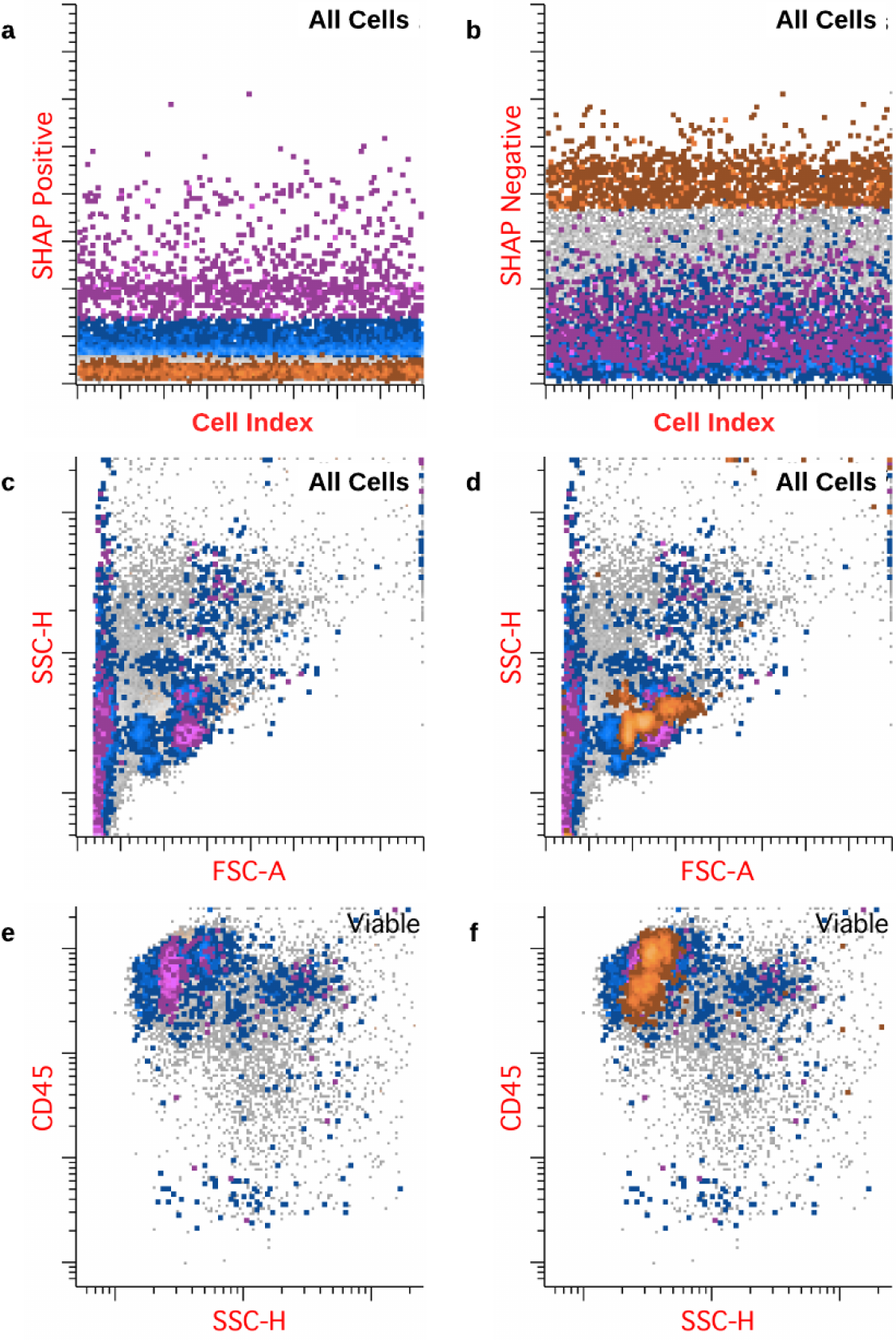
Projection of 2D histogram SHAP values from all 78 2D histograms back to individual cells, plotted using clinical flow cytometry software. The highlighted cells indicate the cell populations that were most impactful in determining the CNN predictions. **a** and **b**: gating of cells by summed SHAP values. Cells highlighted in blue had increased summed positive SHAP values, cells in magenta had very increased summed positive SHAP values, and cells in orange had increased summed negative SHAP values. **c** and **d**: projection of cells along forward scatter and side scatter axes, with (**c**) and without (**d**) superimposed negatively impacting cells highlighted. The cells along the left edge of the plot are consistent with non-viable cells. **e** and **f**: Gated viable cells, with (**e**) and without (**f**) negatively impacting cells highlighted. Most of the clusters of impactful cells are consistent with subpopulations of lymphocytes. Data are from an example case with predicted cHL+ probability > 0.95.

### Integration of SHAP values from all histograms enables visualization of individual cells’ impacts on individual predictions

Using the SHAP values from each of the histogram bins from all 78 2D histograms, we were able to integrate and project the values back to the individual cells in the original raw data files. New FCS^20^ files were created with summed SHAP values for each of the individual cells. These files could then be opened and analyzed using standard flow cytometry analysis software. This results in the ability to define in high detail, in combination with all of the available flow cytometry parameters in the original data and available software tools, the cells of most impact. The example shown in Fig. 7 demonstrates the result, with the most impactful cells highlighted using gating in our usual clinical flow cytometry software. The integration of SHAP values from all 78 2D histograms resulted in sufficient resolution to highlight what appears most consistent with subpopulations of lymphocytes, as demonstrated in the side scatter versus CD45 plot (Fig. 7, parts e and f). Evaluation of CD5 versus CD20 demonstrates the most impactful lymphocytes are primarily T cells. The impactful cells also include many that would normally be classified as non-viable cells and excluded from evaluation in the manual gating process; it is thus likely that the ratio of non-viable to viable cells plays a significant role in the model predictions. Approximately 20 Hodgkin cells were identified through our normal manual gating process, yet none of the Hodgkin cells were found to be as impactful in the classification as the lymphocytes subgroups and non-viable cells, per our summed SHAP values analysis. The evaluation of T cell subpopulations is in fact a part of our normal manual gating analysis, notably with evaluation for overexpression of CD7^21^; however, the necessary antibodies (CD7, in particular) are part of a separately run antibody panel. It is conceivable that these impactful features might eventually be learned and used by pathologists (e.g., via non-viable cells and lymphocyte ratio scores). It is interesting to note that the SHAP summation approach presented here highlights lymphocyte subpopulations as being important, despite the lack of some antibodies that are more specific to lymphocytes and antibody choices primarily chosen to identify Hodgkin cells.

## Discussion

We present a deep learning, ensemble-based approach to detect classic Hodgkin lymphoma using flow cytometry data. Currently pathologists visually interpret two-dimensional histograms, followed by various gating strategies to highlight cells of interest. We applied visual machine learning techniques, namely convolutional neural networks (CNNs), in order to simulate to some degree the process used by pathologists. CNNs were also chosen because they might better preserve local context with possible improvement in results compared to prior techniques^8^. Our deep learning approach does perform well in comparison with prior results of random forest and support vector machine approaches^8^. Despite our approach being agnostic to the important cell populations (viz., we use non-gated 2D histograms), the algorithm is able to learn useful features, and some of the most impactful 2D histograms correlate with clinical experience in using the Hodgkin lymphoma flow cytometry assay. Perhaps of more interest, however, is the fact that the model is intuitively easy for pathologists, researchers, and, potentially, regulators to understand since its architecture and outputs mirror those of the standard manual method. Other flow cytometry machine learning techniques often include some sort of principal component analysis or clustering followed by a classifier^22–24^. They are very successful^25^, but identification of cells of interest in principal components space and other representations is not intuitive for most pathologists. In our EnsembleCNN approach, the architecture is amenable to explainability algorithms and impactfulness measures, including SHAP values^12,13^. Calculation of SHAP values for bins within 2D histograms is potentially more enlightening to hematopathologists since the graphical output can be compared directly with histograms that hematopathologists and researchers are already accustomed to interpreting.

Perhaps of most interest, using SHAP values and a novel summation approach that integrates SHAP values from all of the component 2D histograms, we are able to visually highlight cell populations that are most impactful in making the classification predictions, both as an average for many cases and for individual cases. We can visualize the thus highlighted cells using flow cytometry software (Fig. 7). Doing so allows us to directly visualize which cell populations are most impactful in the EnsembleCNN classifier’s individual predictions. We can directly compare those cells with cell populations identified using standard manual gating approaches and hematopathologist interpretations. In the example presented in Fig. 7, which was strongly predicted to be positive for cHL by the EnsembleCNN classifier (with concordant annotation label), the populations of most impact were non-viable cells and T lymphocytes. As increased numbers of apoptotic cells and altered T cell populations are expected to be found in cHL, the findings are consistent with the currently understood biology^26,27,21,28^. Given the overall agnostic approach of the combined CNN ensemble and SHAP value summation methods, this approach might be useful in the discovery of previously unrecognized cell subpopulations in flow cytometry applications, in cHL or otherwise.

Of note, in the example case (Fig. 7) none of the ∼20 Hodgkin cells identified by manual gating (of 53,591 total flow cytometer events) were identified as very impactful by summed SHAP scores, suggesting that the approach is insensitive to very small cell populations (though larger non-viable and lymphocyte populations were impactful). Use of 2D histograms of gated subpopulations (e.g., viable cells, CD30+ cells, etc.) is expected to increase model accuracy and detection of these small populations as the signal to background ratio is increased. Due to the ensemble architecture, it is simple to add additional features, including gated histograms, to the classifier. The features used for prediction in this report are 2D histograms derived from a single antibody panel (targeted for cHL identification); however, in regular clinical practice, multiple antibody panels are applied to the same specimen. These additional data could easily be used to train additional CNNs that could be added to the ensemble, since each CNN in the ensemble is trained independently. Additional features might also include 3D histograms and other higher dimensional data, as well as FlowSOM^24^ and UMAP^22^ data representations. Insights provided by SHAP values could also guide selection of new features/gated histograms for inclusion.

Limitations of the approach include the training time required to train all CNNs within the EnsembleCNN classifier, which is substantially more than that required for support vector machine and random forest approaches. Since the training time is a one-time cost, the impact on daily use is much less. In the method’s current implementation, the 2D histograms contain data for all cells rather than gated subpopulations; inclusion of gated populations will allow more direct comparison with regular clinical flow cytometry use and identification of known important cell populations with small numbers present (e.g., rare Hodgkin cells). Introduction of gated plots that only contain data for specific subpopulations may require modification of our SHAP values summation approach since the gated plots will cause overrepresentation of those cells in comparison to the rest of the data, for *de novo* cell population discovery. Our SHAP values summation approach, as is, can only be applied to cells that are present within the same antibody panel. Overcoming these obstacles is the object of further work.

In summary, the EnsembleCNN classifier presented here is intuitively easy for hematopathologists to understand, can easily be extended to incorporate additional data features (e.g., gated histograms and additional antibody panels), and is amenable to explainability algorithms and metrics, including SHAP values. Our method for projecting SHAP values back to the original single-cell data allows direct evaluation by, and comparison with, standard flow cytometry analysis. This provides further understanding of the important cell populations and how the model makes a prediction. Use of the approach can potentially save pathologists time by prepopulating clinical reports and pre-highlighting the cells of most interest for pathologist review. This may also be useful in discovery of previously unrecognized important cell populations or other cell population alterations in disease pathogenesis.

## Data Availability

Algorithm code and updates will be made available at http://github.com/SimonsonLab/EnsembleCNN.

## Acknowledgments

We thank the University of Washington hematopathology staff for technical help and support of this work. We thank Patrick Mathias (University of Washington) for review of the manuscript prior to submission.

