## Supplemental Information for "*De novo* identification and visualization of important cell populations for classic Hodgkin lymphoma using flow cytometry and machine learning"

**Supplementary Text**

We were also interested in knowing whether passing the outputs from the penultimate layers of the CNNs (as opposed to the prediction values passed from the final layers; see Fig. 1) as inputs to the random forest classifier would improve the accuracy of the model; however, no significant difference in performance measures was seen, as this other approach resulted in an accuracy of 88.6%, a precision of 82.7%, a recall of 69.4%, an F1 of 75.4%, and an AUROC of 0.93.

Since columns of the 2D dendrogram (Fig. 2) with higher/lower variance are hypothesized to have more/less discriminatory power, the variance was calculated for each column. The five CNNs with most variance include CNNs trained on SSC-A/CD71, SSC-A/CD15, SSC-A/CD64, SSC-H/CD64, and CD20/CD15 2D histograms. Given the common findings among the two approaches, it was also surmised that higher variance in the CNN output correlates with increased discriminatory power/impactfulness (but is much less informative than using the SHAP values).

**Supplemental Tables**

Supplemental Table ST1. Confusion matrix for CNN ensemble classifier.

| CNN ensemble classifier | Predicted - | Predicted + |
|---|---|---|
| True - | 174 | 9 |
| True + | 20 | 42 |

**Supplemental Figures**

a

b

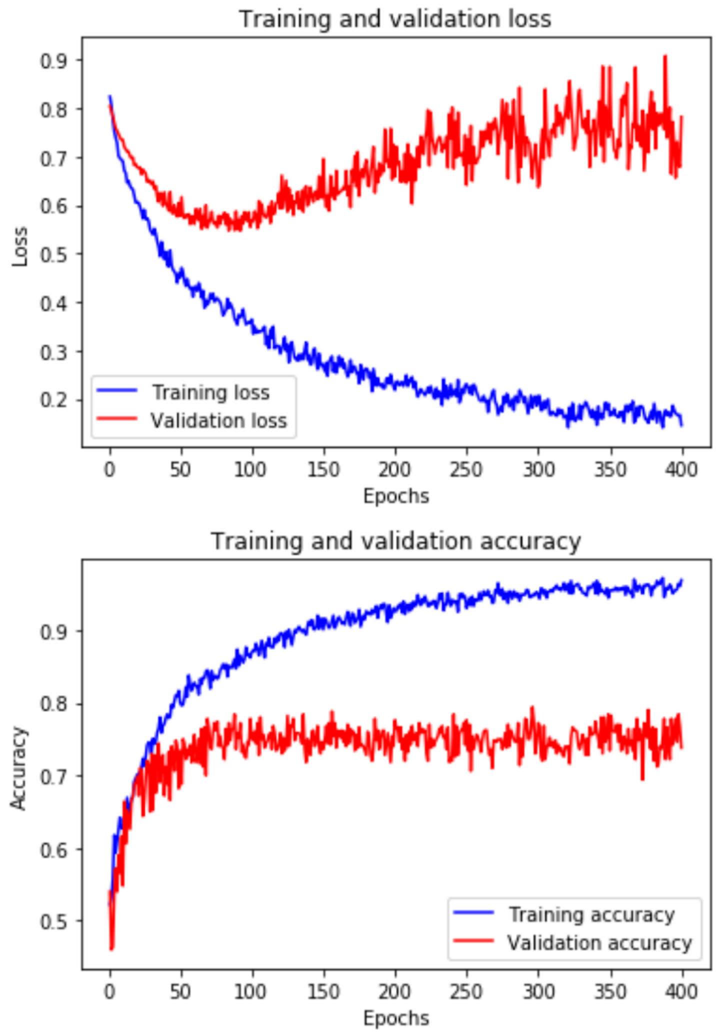


Fig. S1. Typical training loss (**a**) and accuracy (**b**) plots for a single CNN model.

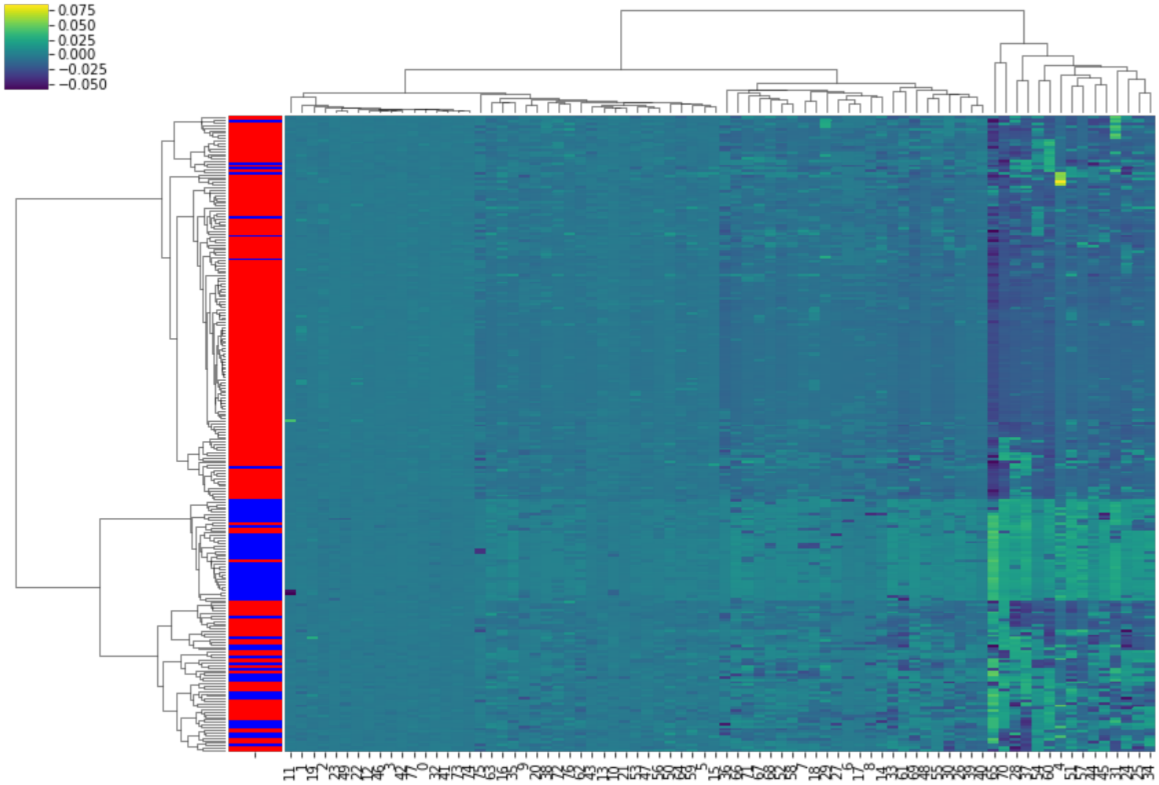


**Fig. S2.** Hierarchical dendrogram of SHAP values for each CNN prediction for each case in test data set. Rows correspond to individual flow cytometry specimens, and each has 78 SHAP values, one for each of the CNNs' predictions. Row labels (left side of plot) correspond to data labels (blue = positive for cHL, red = negative by annotation). The plot demonstrates how the CNN predictions for each case impacted the final prediction (lighter colors corresponding to positive impact while darker have negative impact on predicting cHL positivity). The cases annotated as positive have generally increasing SHAP values in reading the graph from left to right. Constructed using Python seaborn module with metric="euclidian" and method="ward". See also Fig. 4 and Fig. S3.

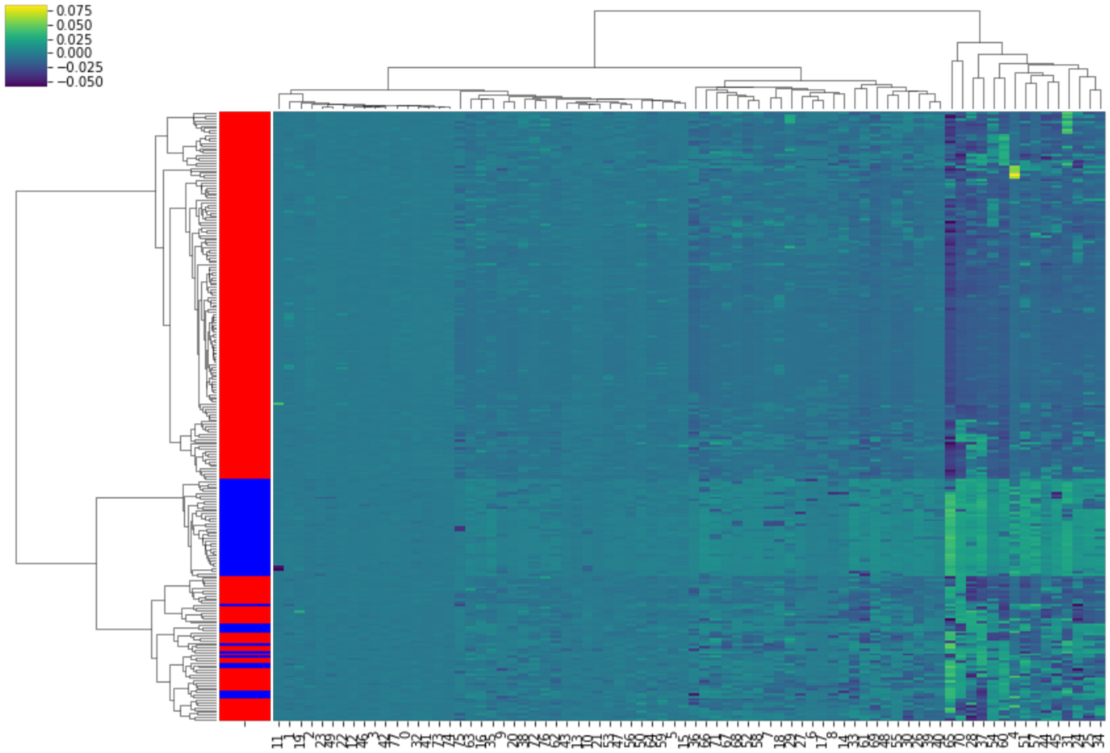


**Fig. S3.** Hierarchical dendrogram of SHAP values for each CNN prediction for each case in test data set. Rows correspond to individual flow cytometry specimens, and each has 78 SHAP values, one for each of the CNNs' predictions. Row labels (left side of plot) correspond to CNN ensemble classifier final predictions (blue = positive for cHL, red = negative by annotation). The plot demonstrates how the CNN predictions for each case impacted the final prediction (lighter colors corresponding to positive impact while darker have negative impact on predicting cHL positivity). The cases predicted to be positive have generally increasing SHAP values in reading the graph from left to right. Constructed using Python seaborn module with metric="euclidian" and method="ward". See also Fig. 4 and Fig. S2.
